# Overexpression of transposable elements is associated with immune overdrive and poor clinical outcome in colorectal cancer patients

**DOI:** 10.1101/2020.07.14.20129031

**Authors:** Xiaoqiang Zhu, Hu Fang, Kornelia Gladysz, Jayne A. Barbour, Jason W. H. Wong

## Abstract

**Objective:** The immune system plays a key role in protecting against cancer. Increased immune infiltration in tumor tissue is usually associated with improved clinical outcome, but in colorectal cancer (CRC), excessive immune infiltration has also been shown to lead to worst prognosis. The factors underlying this immune overdrive phenotype remains unknown.

**Design:** Using RNA sequencing data from The Cancer Genome Atlas, the expression of over 1,000 transposable element (TE) subfamilies were quantified using the “REdiscoverTE” pipeline. Candidate prognostic and immunogenic TEs were screened by survival and correlation analysis, respectively. Based on these candidates, a TE expression score was developed and CRC patients were clustered using the “kaps” algorithm.

**Results:** In CRC, we found that the TE expression score stratified patients into four clusters each with distinctive prognosis. Those with the highest TE expression were associated with immune overdrive and had the poorest outcomes. Importantly, this association was independent of microsatellite instability status and tumor mutation burden. To link TE overexpression to the immune overdrive phenotype, we showed that cell lines treated with DNA methyltransferase inhibitors also had a high TE expression score and activation of cellular innate immune response pathways. Finally, a pan-cancer survey of TE expression identified a subset of kidney renal clear cell carcinoma with a similar adverse immune overdrive phenotype with poor prognosis.

**Conclusion:** Our findings reveal that TE expression is associated with immune overdrive in cancer and is an independent predictor of immune infiltration and prognosis in CRC patients.

**What is already known about this subject?:**

- Cancers with high immune infiltration generally have better prognosis, but it is unknown why a subset of colorectal cancers (CRC) with high immune infiltration have the poorest outcomes.
- Transposable element (TE) expression has been shown to be strongly associated with immune infiltration in cancers but its role in patient prognosis is unclear.
- TEs can be reactivated by DNA hypomethylation in cancers, resulting in immune response via viral mimicry.

**What are the new findings?:**

- A TE expression score has been developed that is predicative of prognosis in CRC patients where those who have the highest TE score show an immune overdrive phenotype and have the worst prognosis.
- The TE expression score predicts prognostic and immune infiltration independent of microsatellite instability and tumor mutation burden (TMB).
- Immune response pathways and infiltrate profiles of high TE expression CRC recapitulates those of DNA methyltransferase inhibitor treated cells where TEs are reactivated, suggesting that TE overexpression may drive immune infiltration in CRC.
- A pan-cancer analysis found that kidney renal clear cell carcinoma shares are a similar TE expression associated immune overdrive phenotype with adverse prognosis.

**How might it impact on clinical practice in the foreseeable future?:**

- Our work highlights the importance of TE expression in evaluating CRC patient prognosis.
- The association of TE expression with the immune overdrive phenotype independent of MSI and TMB status suggests that by considering TE expression, there may be new opportunities to identify MSS CRC patients for immunotherapy and develop new strategies to harness TE driven immune response.

## Introduction

Almost half of the human genome is comprised of transposable elements (TEs). They are also known as “jumping genes” with the ability to move or make copies of themselves to other locations in the genome. TEs are divided into class 1 retrotransposons and class 2 DNA transposons, both of which are further subclassified into subclasses, superfamilies and over 1,000 subfamilies [1]. TEs are mainly epigenetically silenced in normal tissues [2] but can become reactivated due to DNA hypomethylation in cancers [3], resulting in the transcription of retrotransposons into RNA or direct transposition of DNA transposons. One potential consequence of the reactivation of TEs is to stimulate the immune system via viral mimicry [4, 5]. For instance, the human endogenous retrovirus (hERV) was shown to be reactivated by DNA methyltransferase (DNMT) inhibitors, which was accompanied by the up-regulation of viral defense pathways in ovarian [6] and colorectal [4] cancer cells. Recently, it has been shown that some TEs such as hERVs can also serve as tumor antigen signals [5]. These observations have demonstrated the critical roles of TEs in anti-tumor immunity. Nonetheless, how TE expression influences cancer progression and clinical outcome remains unclear.

Patients whose cancer have higher immune cell infiltration tend to have better prognosis. For instance, Immunoscore has been developed based on the density of CD3+ and cytotoxic CD8+ T cells in the tumor and the invasive margin in colorectal cancer (CRC) [7], and has been shown to have prognostic value superior to American Joint Committee on Cancer (AJCC) stage classification [8]. Intriguingly, a recent study identified a high-risk subgroup of CRC patients with high tumor immune infiltration as indicated by high CD8A and CD274 gene expression [9]. Termed “immune overdrive” signature, the subgroup of patients with this signature included both microsatellite instability (MSI) and stable (MSS) status, increased TGF-β activation and overexpression of immune response and checkpoint genes. While whether such patients are likely to benefit from immune checkpoint inhibitors therapy remain to be evaluated, the underlying factor behind this phenotype remains unknown.

Given the recent evidence for the role of TEs in triggering cancer immune response, in this study, we report that TE overexpression is a key factor associated with the immune microenvironment in CRC. Importantly, TE overexpression is predicative of an immune overdrive signature that is associated with poor prognosis in a MSI and tumour mutation burden (TMB) independent manner. We show that the TE expression maybe induced by genome-wide hypomethyation, which is common in CpG island methylator phenotype (CIMP) cancers. Finally, we show that the phenomenon of TE expression-immune overdrive is not restricted to CRC but also present in other cancer types.

## Materials and Methods

### Quantification of transposable element expression

We used the REdiscoverTE pipeline to quantify TE subfamily expression based on RNA sequencing data as described by Kong et al [10]. A detailed description of the pipeline is described in the Supplementary Methods.

### Search for TEs associated with survival and immune activation

To select out the potential TEs associated with survival, we performed univariable Cox regression analysis on four endpoints for survival analysis in CRC cohort including overall survival (OS), DSS (disease specific survival), DFI (disease-free interval) and PFI (progression-free interval) [11]. To estimate the correlation of TEs with immune activity, we included a total of 29 immune activation indices. See Supplementary Methods for further details.

### Patient and Public Involvement

No patient or members of the public were involved in the design of this study.

## Results

### Identification of TEs associated with survival and immune activation in CRC

To establish whether TEs are associated with CRC prognosis and immune activation (summarised in Fig. S1A), we first quantified TE expression by applying the recently developed “REdiscoverTE” pipeline [10] on TCGA CRC RNA sequencing data which has been shown to outperform three existing methods by the original paper including Repenrich [12], SalmonTE [13] and the approach used by Rooney et al [14]. In brief, the pipeline quantifies the number of reads mapping to each TE subfamily without uniquely identifying individual instances in the genome. Our downstream analysis was focused on 1,052 TEs subfamilies, which were classified into five classes including long terminal repeats (LTR), DNA, long interspersed nuclear element (LINE), short interspersed nuclear element (SINE) and Retroposon (Fig. S1B). The expression pattern of these five classes is shown in Fig. 1A, indicating that Retroposon and SINE had higher expression followed by LINE while LTR and DNA had lowest expression [10].

**Figure 1.**
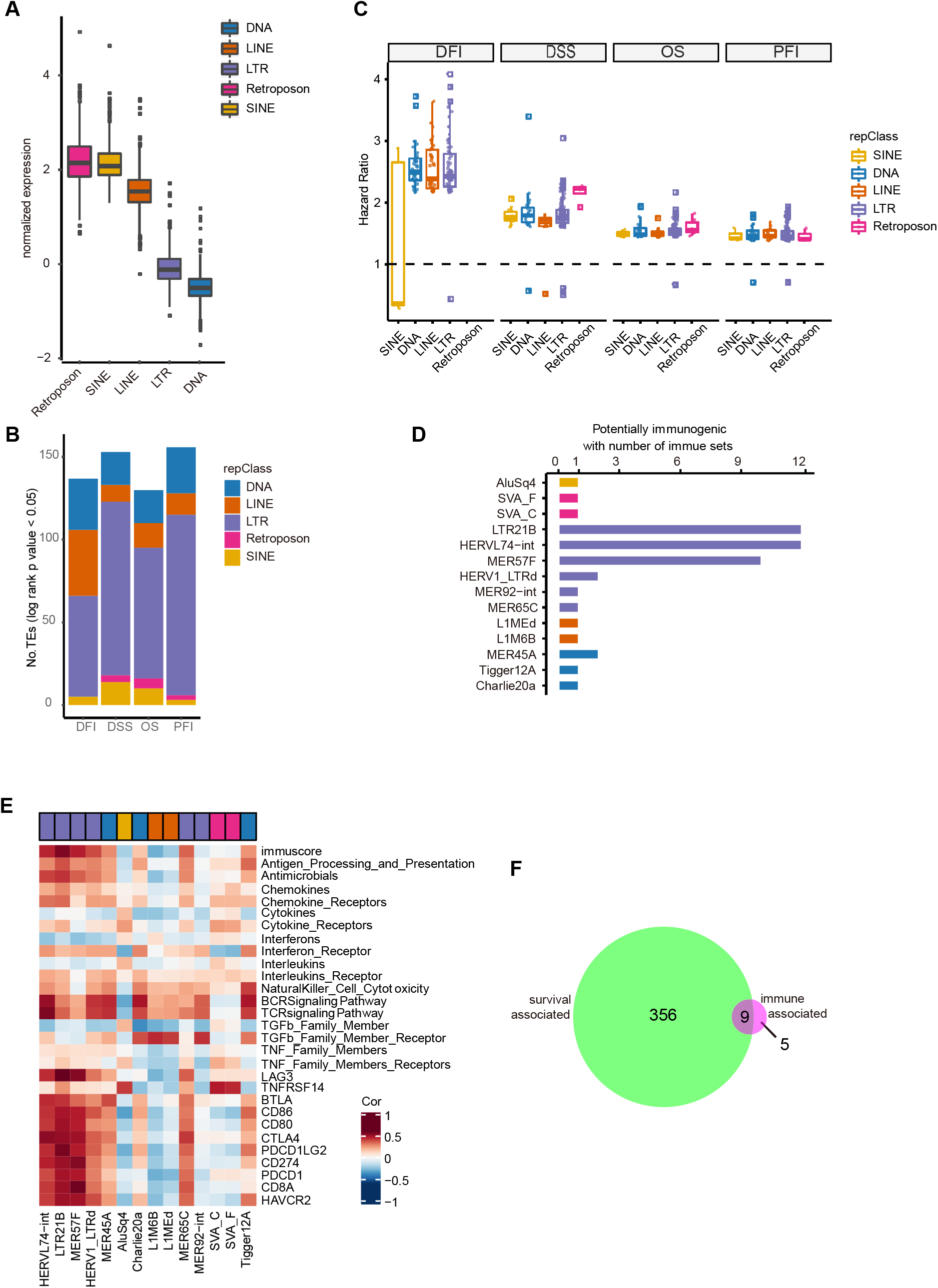
Identification of TEs associated with survival and immune sets in CRC. **(A)** Normalized TE expression pattern at class level including Retroposon, SINE, LINE, LTR, Satellite and DNA. **(B)** Stacked plot showing the number of subfamily TE with significant log rank p value (p < 0.05) for each of the four endpoints including DFI, DSS, OS and PFI. TEs were annotated at class level. **(C)** Distribution of hazard ratios of significant TEs from **(B)** for each of the four endpoints. TEs were annotated at class level. **(D)** Number of immune sets significantly correlated with each TE (Cor ≥ 0.4, p < 0.0001). **(E)** Spearman’s correlation between candidate TE expression (n=14) and 29 immune sets. Heatmap colors indicate the correlation coefficient. **(F)** Venn diagram showing 9 TEs overlapped between candidate prognostic TEs and immunogenic TEs.

To identify prognostic TEs, we performed survival analysis on each TE in terms of four endpoints, respectively, including OS, DSS, DFI and PFI. 365 candidate TEs had survival differences for at least one endpoint (Fig. 1B, Table. S1) with seven significant in all the four endpoints (Fig. S1C-G). Using a permutation test (see Methods), we estimated the average of false discover rate (FDR) to be 1.35% (Fig. S1H). Interestingly, almost all of the hazard ratio of the candidate TEs were greater than one, indicating that higher TE expression generally contributed to worse survival (Fig. 1C). Similar analysis was performed at family and class level, respectively. Retroposon showed significant differences in terms of three endpoints (Fig. S1I and J). Further multivariable Cox regression analysis for these retroposons suggested five to be independent predictors of survival for at least one endpoint (Fig. S1K-M).

Next, we correlated individual TEs with 29 immune indices to identify immunogenic TEs (see methods, Fig. S1N) and found that 14 out of the 1,052 TEs had significant positive correlation with at least one immune index (Spearman’s correlation ≥ 0.4, p < 0.0001, Fig. 1D and E). The FDR of the significant immune-TE associations was estimated to be 0.7% (Fig. S1O).

To integrate the results of the prognostic and immune TE associations, we overlapped the 365 survival and the 14 immune positively correlated TEs. Nine overlapping TEs were identified and used for further exploration (Fig. 1F). All nine TEs were overexpressed compared with normal tissue in at least one cancer type with seven being statistically significant [10] (Fig. S1P). Thus, these TEs can be generally considered to be overexpressed in cancer samples.

### Generation of CRC subtypes based on TE score and clinical outcome

Based on the nine TEs identified above, we first explored the relationship between their expression, finding that most were positively correlated with each other (Fig. S2A). As such, to generate a combined TE score, we averaged the normalised TE expression across CRC samples. We applied the kaps algorithm [15] to the normalised TE score with OS (see Supplementary Methods, Fig. S2B-G) and identified four-risk groups termed TE cluster 1 to 4 with increasing TE score (Fig. 2A). There were significant differences among these four clusters in terms of some molecular features (Fig. 2B-E, Fig. S2H, Table. S2). Specifically, cluster 4 showed higher fraction of MSI samples (33%) but lowest for cluster 1 (11%) (P = 0.0001, Fig. 2B). Cluster 4 also had more samples with CpG island methylator phenotype (CIMP) (30%) (P = 0.0228, Fig. 2C). We also observed some overlaps between TE clusters and other two molecular subtypes including consensus molecular subtype (CMS) [16] (Fig. 2D) and immune subtypes [17] (Fig. 2E). Lastly, there were significant survival differences among these four clusters (log-rank P = 0.0035 for OS, P = 0.011 for DSS, P = 0.12 for DFI and P = 0.022 for PFI, Fig. 2F-I). Generally, cluster 4 showed worse survival while cluster 3 showed the most favorable outcomes (cluster 4 versus 3 with HR = 2.36, 95%CI = 0.96-5.80, P = 0.05 for OS, HR = 3.99, 95%CI =1.12-14.16, P = 0.02 for DSS and HR = 3.05, 95%CI = 1.34-6.94, P = 0.005 for PFI). Notably, cluster 4 had a very poor survival rate after relapse while cluster 3 had superior survival rate after relapse (Fig. 2I).

**Figure 2.**
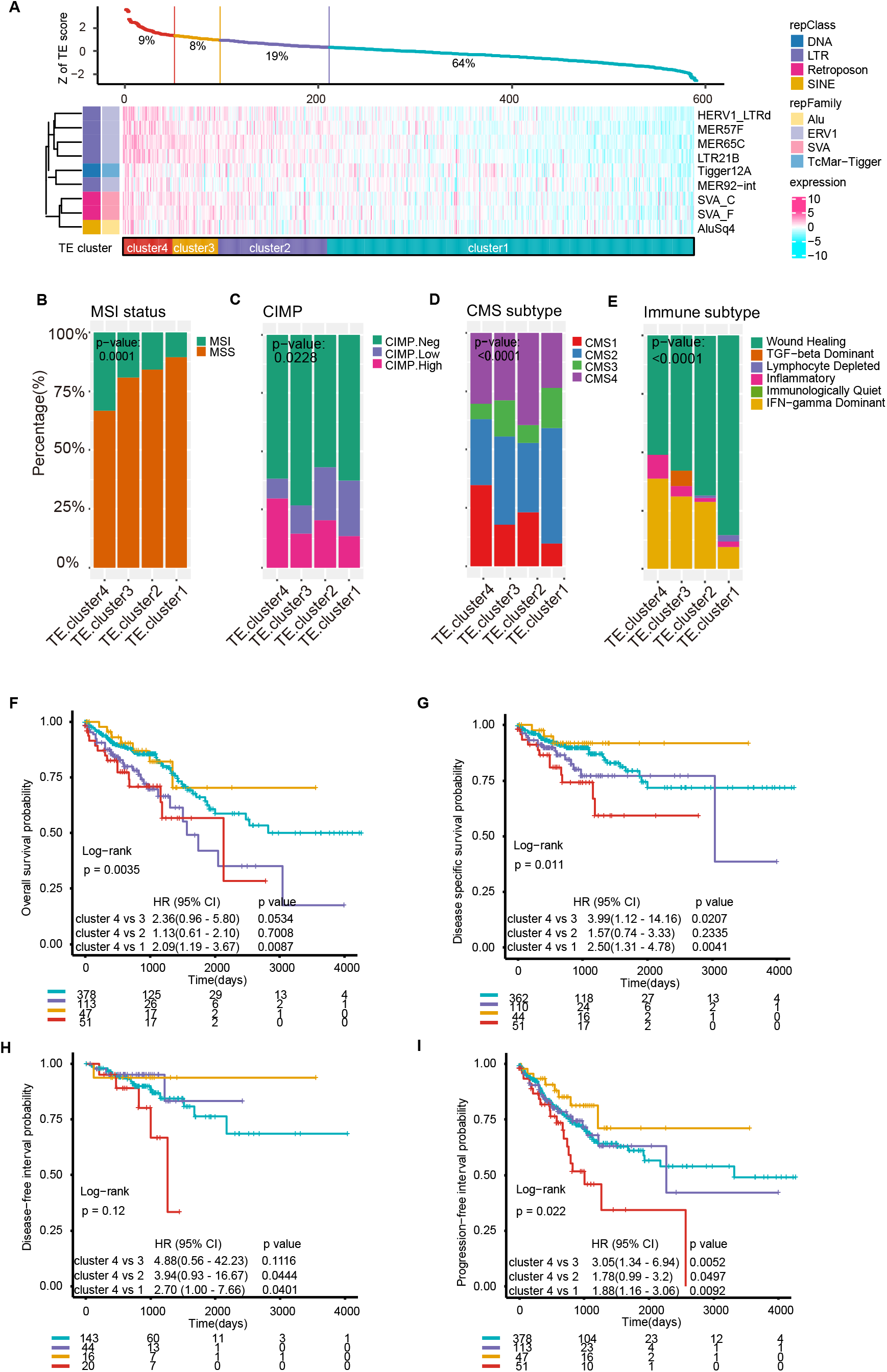
Generation of TE score-based CRC clusters and comparison of molecular association. **(A)** Top scatter plot showed the TE score in decreasing order from left to right. Bottom heatmap displayed the expression profiles of the 9 TEs across four TE clusters as ordered by the TE score. Each TE was annotated at family and class level, respectively. **(B-E)** Stacked plots showing the fractions of molecular features across four TE clusters including MSI status **(B)**, CIMP **(C)**, CMS subtypes **(D)** and immune subtypes **(E)**. **(F-I)** Prognostic value of four TE clusters with Kaplan-Meier survival analysis for OS (n = 589) **(F)**, DSS (n = 567) **(G)**, DFI (n = 223) **(H)** and PFI (n = 589) **(I)**. The hazard ratios (HR) and 95% confidence intervals (CIs) for pairwise comparisons in univariable analyses (log-rank test) are displayed in each Kaplan-Meier plot. Numbers below the x-axes represent the number of patients at risk at the selected time points. The tick marks on the Kaplan-Meier curves indicated the censored patients. MSI, microsatellite instability; CIMP, CpG island methylator phenotype; CMS, consensus molecular subtype.

### TE score is a prognostic and immune infiltration predictor independent of MSI and tumor mutation burden

In CRC, it is well established that patients with tumors that are MSI or have high TMB generally have better prognosis [18, 19]. As TE cluster 4 are slightly enriched with MSI tumors compared with other groups (33% versus 19%, 16% and 11% in clusters 3, 2 and 1 respectively), we sought to determine whether TE score is an independent predictor of prognosis and immune infiltration. After adjusting for clinical factors, cluster 4 remained an independent prognostic variable for three endpoints with HR against cluster 3 of 3.98 (95%CI: 1.09-14.57, P = 0.037) for OS, 9.52 (95%CI: 1.18-76.54, P = 0.034) for DSS and 2.79 (95%CI:1.07-7.31, P = 0.036) for PFI (Fig. 3A-C). Subgroup analysis indicated that the four cluster were well separated especially in MSI samples (Fig. 3D and E). Moreover, TMB (defined as non-silent mutations per megabase, Mb) [20] correlated poorly with TE score (r = 0.13, Fig. 3F) and displayed no difference among the TE clusters (Fig. 3G).

**Figure 3.**
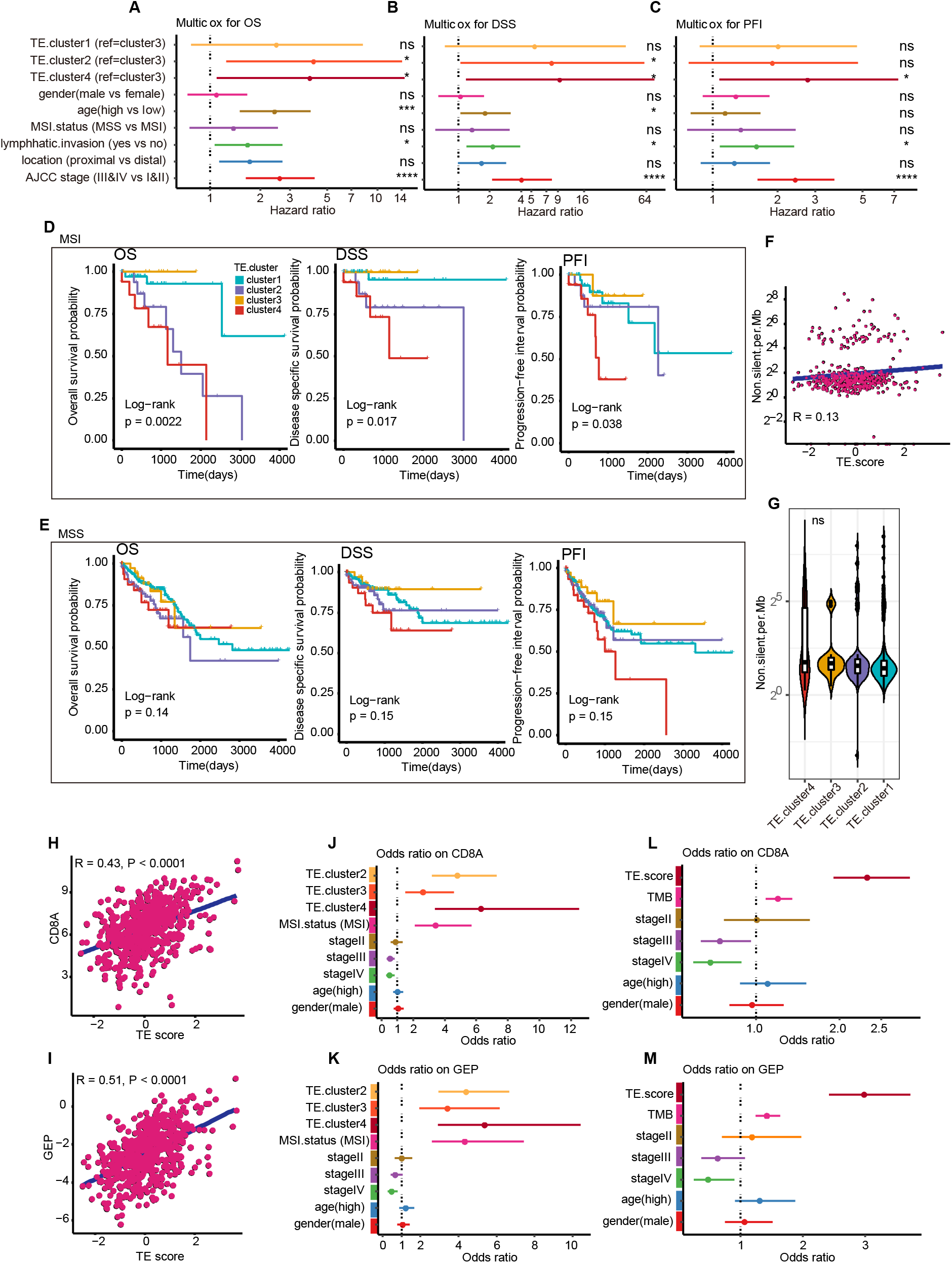
Prognostic value of TE cluster and immune infiltration prediction. **(A-C)** Forest plots showing multivariable Cox regression analysis of TE cluster adjusted by clinical features for OS **(A)**, DSS **(B)** and PFI **(C)**. All variables were set as categorial variable. Samples with age < 65 was set as age low group and ≥ 65 for high group. Solid dots represent the HR of death and open-ended horizontal lines represent the 95 % confidence intervals (CIs). All p-values were calculated using Cox proportional hazards analysis (ns: p > 0.05, *: p <= 0.05, **: p <= 0.01, ***: p <= 0.001, ****: p <= 0.0001). **(D-E)** Prognostic value of four TE clusters with Kaplan-Meier survival analysis in two subgroups separated by MSI status (MSI in D, MSS in E) for three endpoints, respectively. DFI was excluded because of non-comparable sample size among TE clusters. P-value was calculated using log-rank test. Numbers below the x-axes represent the number of patients at risk at the selected time points. The tick marks on the Kaplan-Meier curves indicate the censored patients. **(F)** Spearman’s correlation between normalized TE score and non-silent mutation per Mb. **(G)** Violin plot comparing non-silent mutation per Mb among TE clusters (n.s.: p > 0.05). **(H-I)** Spearman’s correlation between normalized TE score and CD8A expression **(H)** and GEP **(I)**, respectively. **(J-K)** Forest plots showing the odds ratio indicating immune infiltration determined by CD8A expression **(J)** and GEP **(K)** using multinomial logistic regression analysis adjusted by MSI status. **(L-M)** Forest plots showing the odds ratio indicating immune infiltration determined by CD8A expression **(L)** and GEP **(M)** using multinomial logistic regression analysis adjusted by TMB. Solid dots represent the adjusted OR and open-ended horizontal lines represent the 95 % confidence intervals (CIs). OR to the right of dashed line (where OR = 1) indicates higher odds of immune infiltration while OR to the left of the dashed line indicates lower odds of immune infiltration.

To test if the TE clusters can independently predict immune infiltration, we used CD8A, a maker of CD8+ T cells and a T cell-inflamed gene expression profile (GEP) [21]. Firstly, we observed strong correlations between TE score with CD8A (r = 0.43, Fig. 3H) and GEP (r = 0.51, Fig. 3I), respectively. Multinomial logistic regression analyses demonstrated that cluster 4 was a significant predictor for CD8A and GEP and notably displayed the highest odds ratio (OR) of 6.3 for CD8A (Fig. 3J) and 5.4 for GEP (Fig. 3K). Furthermore, TE score showed much higher OR than TMB with 2.3 versus 1.3 for CD8A (Fig. 3L) and 3.0 versus 1.4 for GEP (Fig. 3M). Our results demonstrated that TE clusters and TE score are predicative of immune infiltration and prognosis independent of MSI and TMB.

### Immune overdrive is associated with TE score and expression

To further confirm that the TE expression score is associated with immune activation, we examined the differences of tumor immune microenvironment (TME) among TE clusters. Both GSVA and MCPcounter analyses indicated that cluster 4 displayed higher fractions of most immune cell types especially for T cells, macrophages and dendritic cells, while cluster 1 showed a lack of immune infiltration (Table. S3, Fig. 4A, Fig. S3A). Further, cluster 4 displayed the highest gene signatures of immune infiltration followed by cluster 3, 2 and lowest for cluster 1 (Fig. 4B). These signatures included T cell, lymphocyte, leukocyte infiltration signatures, hot tumor signature and tumor associated macrophage (TAM) ratio. Cluster 4 also displayed highest expression of T helper 1 (Th-1) immune response and regulatory genes, MCH I/II molecules, higher IFN-γ response rate and T cell exhaustion status (Fig. 4B, Fig. S3B and C).

**Figure 4.**
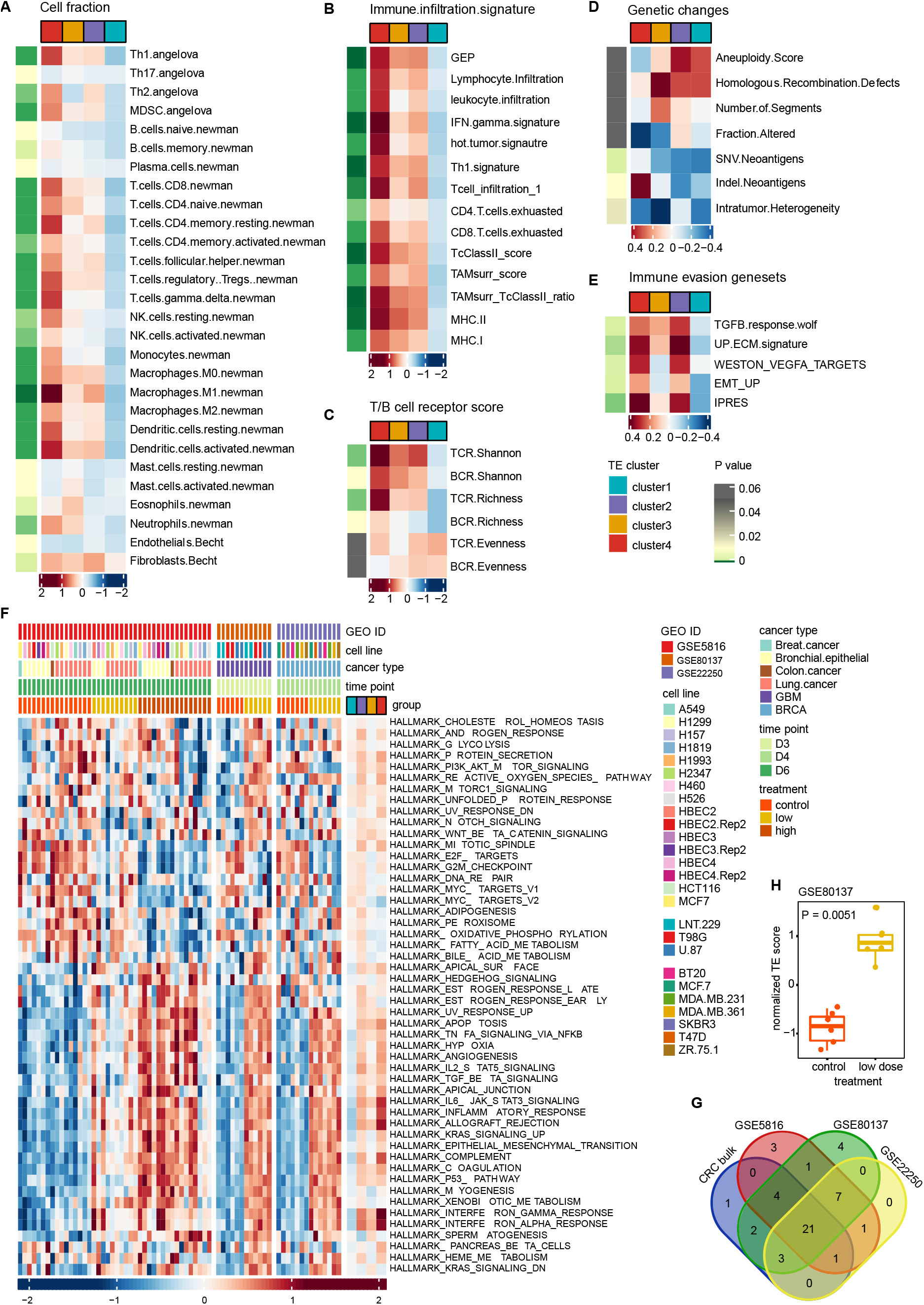
Exploration of immune overdrive phenotype. **(A)** Gene set variation analysis showing fraction of 28 cell types. **(B)** Gene set variation analysis showing immune infiltration signatures. **(C)** TCR/BCR indexes comparison among TE clusters. **(D)** Genetic changes comparison among TE clusters. **(E)** Gene set variation analysis showing immune evasion signatures. P-value for each variable was calculated using Kruskal-Wallis test. For each variable, the median of normalized value in each cluster was shown. **(F)** Heatmap showing 50 hallmark gene sets score based on gene set variation analysis in three 5-aza treated cell line datasets across multiple cell lines and CRC TE clusters. **(G)** Venn diagram showing the overlapped significant pathways among cell line datasets and bulk CRC. **(H)** Comparison of TE score between treated and control groups in GSE80137.

Next, we investigated association of TE clusters with genetic changes but did not find distinctive differences among the clusters except for SNV/Indel neo-antigens (Fig. 4D). Specifically, we investigated associations with hotspot mutations by focusing on 95 CRC specific drivers [22, 23] (see Supplementary Methods, Fig. S3D). Only TP53 (29% in cluster 4 versus 17% in other three clusters) and BRAF (14% in cluster 4 versus 2.4% in other three clusters) displayed significance (P = 0.0001 and P = 0.0378 for TP53 and BRAF respectively). As for the copy number changes shown in Fig. S3E, there was no distinctive differences among TE clusters.

To elucidate the molecular phenotype associated with the TE clusters, we further analyzed dysregulated pathways among TE clusters. We found that cluster 4 and 2 displayed stronger immune evasion-associated signatures including TGF-β response, extracellular matrix (ECM) gene expression, VEGF target, Epithelial–mesenchymal transition (EMT) and innate anti-PD1 resistance (IPRES) signatures (Fig. 4E, Fig. S3F). Furthermore, 32 out of 50 hallmark gene sets were dysregulated among TE clusters and most were upregulated in cluster 4 (Fig. 4F). Similarly, 24 out of 39 drug targetable pathways were significantly different among TE clusters indicating the potential therapy targets for individual clusters such as ALK and PI3K pathways in cluster 4, anti-apoptosis and epidermal growth factor (EGF) signals in cluster 3, plasma membrane signal in cluster 2 and MYC pathway in cluster 1 (Fig. S3G).

Finally, we compared the expression of two markers including CD8A and CD274, which were used to identify immune overdrive by Fakih et al [9]. Our results demonstrated that cluster 4 also displayed higher expression of these two markers (Fig. S3H and I), implying the comparability of immune overdrive identified by TE cluster and these two markers. More importantly, based on the classification of risk groups identified by Fakih et al [9], we found that risk group (IV*) characterized by immune overdrive signature also displayed highest TE score, followed by risk group III* and I/II (Fig. S3J). Together, these results suggest that the immune overdrive phenotype is characterized by the highest TE score (i.e. cluster 4), highest immune infiltration, poorest survival, immune evasion activity (e.g. TGF-β signal), and higher expression of immune response and checkpoint genes.

### Activation of innate immune response in CRC with high TE score recapitulates TE reactivation by DNA methyltransferase inhibitors

To link TE overexpression to the activation of immune response, we compared immune pathway activation in DNMT inhibitors, 5-azacytidine (5-aza) or 5-aza-2’-deoxycytidine (decitabine) treated cancer cell lines and the CRC TE clusters. Generally, activity of hallmark gene sets was consistent in three DNMT inhibitor treated cell lines (Fig. 4F). Interestingly, we observed large overlaps of the significant up-regulated pathways in treated groups of these three data sets and CRC TE cluster 4 (n=21, Fig. 4G, Table. S4). Furthermore, by quantifying the TE expression in decitabine treated GBM cell lines (GSE80137), we found that TE score were highest in the decitabine treated groups compared to control group (Fig. 4H).

As the DNMT inhibitor treated samples are all derived from cell lines, to further confirm that TE expression signals are mainly derived from tumor cells in bulk CRC tissue rather than the TME, we investigated TE expression in a high-depth single cell RNA sequencing (scRNA-seq) breast cancer dataset [24] (Fig. S3K). Importantly, the proportion of reads mapping to TEs was highest in tumor cells (P = 0.0019, Fig. S3L).

### TEs trigger intracellular immune response by viral mimicry

To comprehensively examine pathways driven by TE overexpression in CRC, we applied a weighted correlation network analysis (WGCNA) [25] to find gene modules that were associated with TE score (see Supplementary Methods). Our results suggested that two of the module genes were strongly positively correlated with TE score (r= 0.5 for greenyellow module, r = 0.46 for brown module, Fig. 5A, Fig. S4A-G, Table. S5). There were 39 and 389 genes in greenyellow and brown module, respectively. To determine the function of these modules, we performed GO and KEGG enrichment analysis using these genes (Fig. 5B and C). Genes in the greenyellow module were enriched in innate immune pathways, while the brown module was more enriched in adaptive immune response such as differentiation, migration and activation of immune cells.

**Figure 5.**
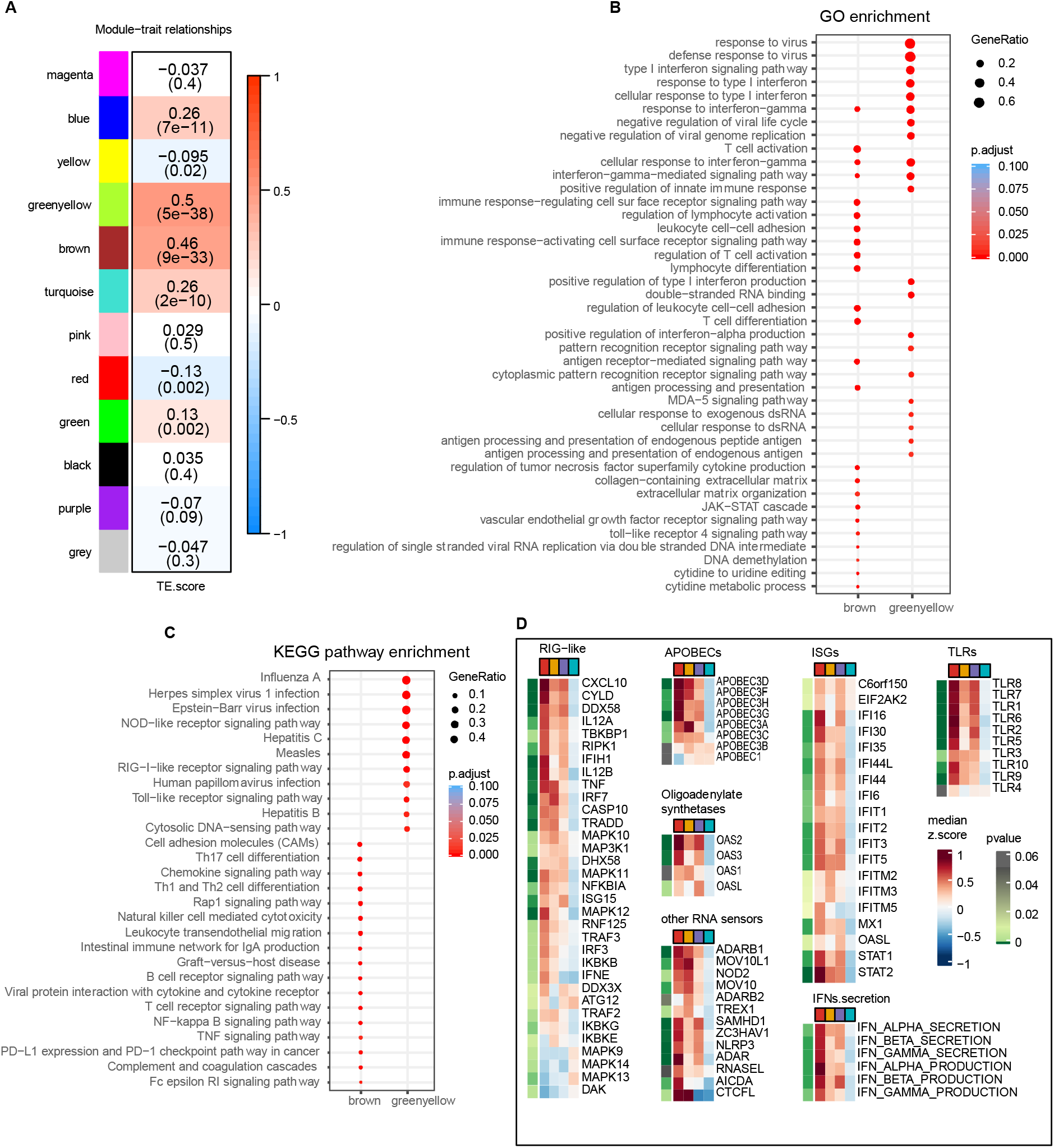
Weighted gene co-expression network analysis (WGCNA) based on TE score. **(A)** WGCNA consensus network modules correlated with TE score. Each row corresponds to a module, column to the TE score, respectively. Each cell contains the corresponding correlation coefficient and p-value. Individual gene modules were marked using different colors. **(B)** GO enrichment analysis of the genes in the brown and greenyellow module, respectively. **(C)** KEGG pathway enrichment analysis of the genes in the brown and greenyellow module, respectively. The size of the circle indicates the ratio of the genes mapped to each pathway. **(D)** Representative expression of genes or signatures involved in immune response and RNA sensor signals including RIG-I-like pathways, APOBECs, Oligoadenylate synthetases, RNA sensors, interferon-stimulated genes (ISGs), interferon secretion process and Toll-like receptors (TLRs). P-value for each variable was calculated using Kruskal-Wallis test. For each variable, the median of normalized value in each cluster was shown.

We then further compared the expression of some critical genes that might be involved in the response to TE reactivation. As expected, most of these genes were highly expressed in cluster 4 followed by cluster 3 and 2 and lowest in cluster 1 (Fig. 5D). Specifically, the secretion and production of IFN α and β were increased in cluster 4 which indicated type I IFN response. Similarly, TE suppressors such as APOBECs, ADAR, NOD2, MOV10, MOV10L1 and CTCFL were also upregulated in cluster 4. It has been shown that CTCFL, a germline-specific transcription factor, functions as suppressor of SVA expression by directly binding to and regulate SVA repeats [26]. These findings support a role for TE overexpression triggering innate immune response in a manner similar to viral invasion.

### Pan-cancer analysis identified immune overdrive phenotype in kidney renal clear cell carcinoma

Finally, to determine if the TE induced immune overdrive phenotype is also present in other cancer types and to ensure that our findings are not cohort specific, we examined TE expression in another 23 cancer types. Firstly, several cancer types showed higher TE score such as kidney renal clear cell carcinoma (KIRC), diffuse large B-cell lymphoma (DLBC) and head and neck squamous cell carcinoma (HNSC) while CRC and adrenocortical carcinoma (ACC) were generally lower (Fig. S5A, Table. S6). By performing univariable Cox regression analysis on the TE score in each cancer type, we found that apart from CRC, KIRC also had significantly increased HR (HR = 1.78, 95%CI = 1.43 - 2.22, P value < 0.0001, Fig. 6A, Table. S7). Further correlation analysis indicated that TE score correlated well with GEP across the cancer types (r = 0.45, Fig. S5B). Individually, the best correlations were observed in SKCM, HNSCC and CESC followed by CRC and PRAD (Fig. 6B).

**Figure 6.**
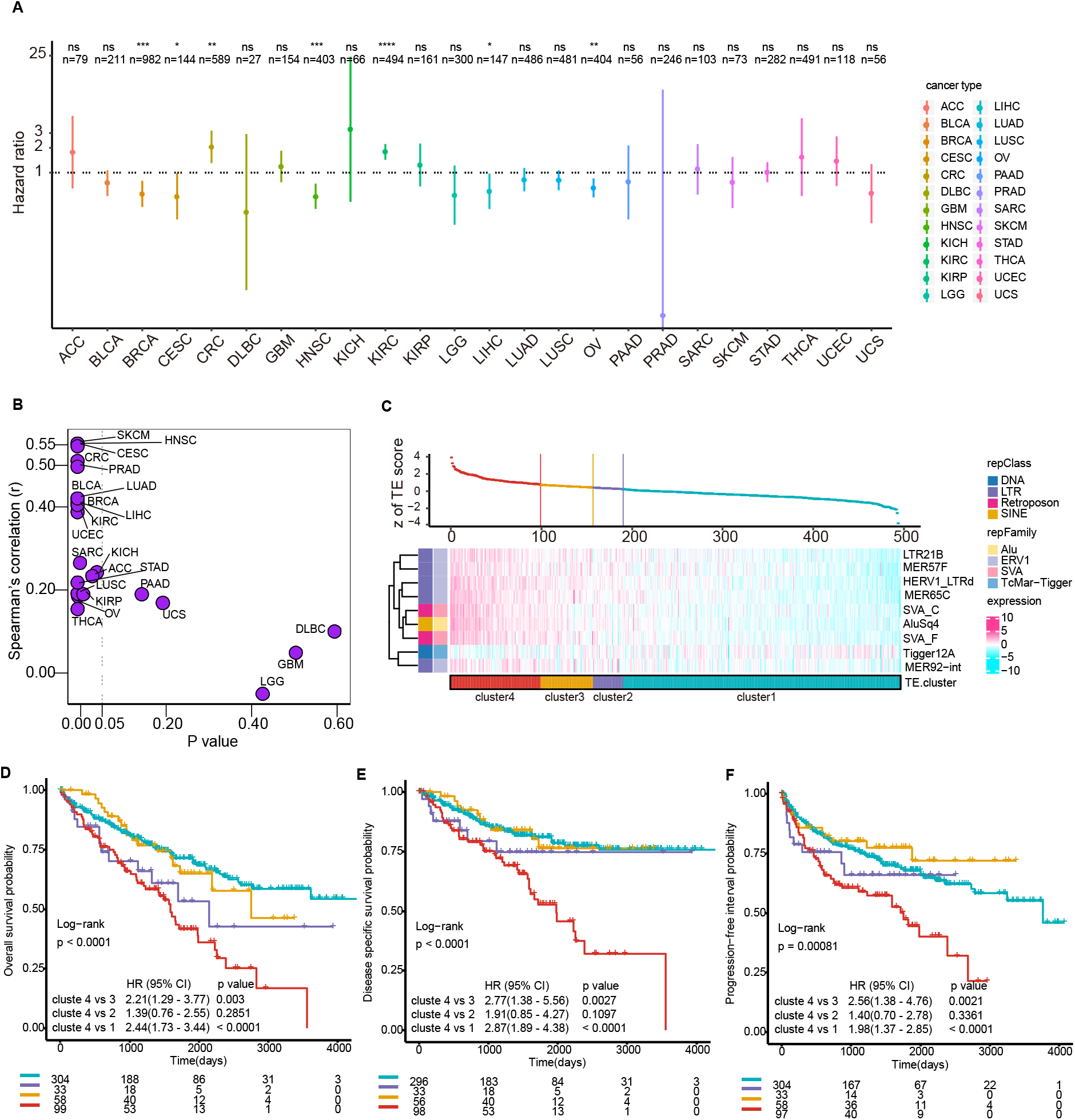
Pan-cancer analysis of TE score and identification overdrive phenotype in KIRC. **(A)** Forest plot showing the univariable Cox regression analysis of OS on TE score across 24 cancer types. Solid dots represent the HR of death and open-ended horizontal lines represent the 95 % CIs. All p-values were calculated using Cox proportional hazards analysis (ns: p > 0.05, *: p <= 0.05, **: p <= 0.01, ***: p <= 0.001, ****: p <= 0.0001). **(B)** Spearman’s correlation between TE score and GEP across 24 cancer types. X-axis indicates p-value and y-axis indicates correlation coefficient. **(C)** Top scatter plot showed the distribution TE score in decreasing order from left to right in KIRC. Bottom heatmap displayed the expression profiles of 9 TEs across four TE clusters as ordered by the TE score. Each TE was annotated at family and class level, respectively. **(D-F)** Prognostic value of four TE clusters with Kaplan-Meier survival analysis in KIRC for OS (n = 494) **(D)**, DSS (n = 483) **(E)** and PFI (n = 492) **(F)**. The hazard ratios (HR) and 95% CIs for pairwise comparisons in univariate analyses (log-rank test) are displayed in each Kaplan-Meier plot. Numbers below the x-axes represent the number of patients at risk at the selected time points. The tick marks on the Kaplan-Meier curves indicate the censored patients.

As some KIRC appeared to exhibit an immune overdrive phenotype, we carried out similar analyses as we had done for CRC. Eight of the nine TEs had similar trends of expression across samples except for Trigger12A (Fig. 6C). As with CRC, we identified four KIRC clusters with differing prognostic outcomes with cluster 4 having the worst outcomes (Fig. 6D-F) and enrichment of molecular subtypes (Fig. S5C). Like CRC, cluster 2 also had relatively short OS while cluster 3 had favorable survival rate after relapse. After adjusting by clinical features, cluster 4 remained significant for all three endpoints and cluster 2 were significant for OS and PFI (Fig. S5D-F).

Consistently, cluster 4 in KIRC also displayed highest immune infiltration and immune evasion phenotypes (Fig. S5G-J). As with CRC, there were no distinctive differences for genetic changes among these TE clusters (Fig. S5K). Finally, most of the genes involved in the response of TE reactivation had higher expression in cluster 4 also showing similar immune regulation patterns with CRC (Fig. S5L-R).

## Discussion

Molecular subtyping based on genomic and transcriptomic data has facilitated improved understanding of molecular features in cancers and has guided targeted strategies in cancer treatment [27]. For instance, MSI is a critical subtype in CRC which has been associated with high immune infiltration (e.g. CD8^+^ T cells) [18] and lower risk of relapse [28]. Generally, cancers with higher immune infiltration had better survival including CRC [7, 17]. However, an immune overdrive phenotype is also observed in CRC, characterized by high immune infiltration but poorest survival [9]. The immune overdrive phenotype can be reproduced using a nine TE expression signature where TE cluster 4 was characterized by the immune overdrive phenotype with the highest TE score, poorest survival but also highest immune infiltration. Our findings highlight the importance of TE expression on not only immune infiltration but also on patient prognosis in CRC.

Compelling evidence indicates the critical roles of reactivated TEs in cancer development and progression resulting from the loss of TEs suppression [12, 29]. Generally, epigenetic regulation, especially DNA methylation and histone modification, are the best-known mechanisms of TE silencing. Indeed, studies have demonstrated that epigenetic alterations could lead to carcinogenesis in which TE reactivation might be a potential secondary cause [30]. The global loss of methylation can lead to TE reactivation and is often accompanied by the hypermethylation of tumor suppressor genes in cancers [31]. For instance, the reactivation of LINE1 caused by DNA hypomethylation has been observed in several cancer types including CRC [32], LIHC [33], and BRCA [34]. It has been shown that DNMT inhibitor treatment could stimulate innate immune response accompanied by TE reactivation including hERVs and other class of TEs [4, 6, 10]. Our analysis based on cell line data showed that DNMT inhibitor treated cells shared many of the dysregulated gene pathways as CRC patients in cluster 4. Although this observation is not direct mechanistic evidence supporting DNA demethylation driven TE overexpression induced immunogenicity in CRC patient samples, it shows a possible relation between TE expression and immune response.

Previous studies have profiled the landscape of TE expression across human tissues and indicated that TE expression is much higher in solid tissues compared with in whole blood [35, 36]. This is in line with our findings based on high depth scRNA-seq dataset of breast cancer that cancer cells are the main contributor of TE expression. Thus, TE expression might reflect a cancer cell intrinsic characteristic [37]. Besides, our analysis revealed that TE score was generally correlated with not only the proportion of reads mapping to TEs but also TE expression at class level in both bulk CRC, KIRC and scRNA-seq dataset (Fig. S6A-D). This indicated that our TE score might reflect global TE expression more generally. Importantly, we found TP53 mutations enriched in cluster 4. TP53 can function to restrain TEs and TP53 mutations may potentially cause reactivation of TEs [38], which may reflect another mechanism for the observed TE overexpression.

Innate immune system is essential for pathogen recognition and initiation of protective immune response through the recognition of pathogen associated molecular patterns (PAMPs) by its pattern recognition receptors (PRRs) [39]. Nucleic acids including RNA and DNA are critical PAMPs especially for viruses. We found that, upon TE overexpression, some important PRRs of innate immune signals were up-regulated. Indeed, TE-derived RNAs are very prevalent and can form dsRNA in the nucleus. Annealing of these hybirds is relaxed by adenosine (A)-to-inosine (I) editing through ADAR or cytidine (C)-to-uridine (U) deamination editing through APOBEC3s [37, 40]. However, the unedited hybirds are prone to bind with RNA sensors and further stimulate downstream immune response by viral mimicry represented by increased IFN response. To date, it remains unknown whether individual classes of TEs are prone to activate different PRRs [41]. TE reactivation can also stimulate immune response through other mechanisms, for example, LTR can function as prompters or enhancers of IFN-stimulated genes [42] and hERVs can a source of antigens [43]. Further investigations will be required to establish the mechanism underlying the association between TE overexpression and immune overdrive.

Currently, measuring and interpreting TE expression is still challenging [44]. Mappability, polymorphisms and transcript identity are three main difficulties for TE transcription study. Although some approaches can also measure TE expression at locus-specific level such as SQuIRE [45] and Telescope [46], the main disadvantage is the low confidence of measurement of youngest TE subfamilies because of less uniquely alignable sequence. Currently, the best way to assess locus-specific TE expression is using full-length RNA-seq data derived from long-read sequencing technologies such as provided by PacBio or Oxford Nanopore [47]. Nevertheless, in this study, we explored the association between TE expression and immune activation at a landscape level. Our goal was not to identify to locus-specific TE expression, and verify their biological function. Thus, measuring the total transcriptional output from a group of related TEs, such as TE family and subfamily levels is sufficient to support our findings.

It has been suggested that CRC with MSI could benefit from immune checkpoint blockade (ICB) therapy. Moreover, epigenetic therapy has been proven to increase tumor immunogenicity and modulate the response to immunotherapy [4]. Thus, there are several potential strategies for the treatment of patients amongst TE clusters. MSI tumors in cluster 4 are prone to relapse, therefore, patients in this cluster might benefit from combined ICB and chemotherapy. Currently, some clinical trials such as ATOMIC, are ongoing with the aim to investigate the efficiency of ICB for MSI-H CRC. Our findings indicated that patients with highest TE score might have higher risk of recurrence and benefit from ICB. In addition, as we showed that TE clusters is predicative of prognosis independent of MSI status and TMB, MSS patients with high TE expression score may also benefit ICB. Furthermore, our results found that activation of TGF-β, ALK, PI3K pathways were enriched in cluster 4. Therefore, these specific pathways might be useful as targets for therapy in combination with ICB. Finally, epigenetic therapy combining with ICB might be suitable for patients in cluster 1 and 2 with relative lower expression TE. More studies and clinical trials will be needed to confirm these strategies.

In conclusion, our results highlight the importance of TE expression in evaluating CRC patient prognosis. The association of TE expression with the immune overdrive phenotype independent of MSI and TMB status suggests that by considering TE expression, there may be new opportunities to identify MSS CRC patients for ICB and develop new strategies to harness TE driven immune response.

## Data Availability

TCGA CRC RNA sequencing data were directly analysed on Cancer Genomics Cloud using a custom pipeline [http://www.cancergenomicscloud.org/]. The processed TE expression for pan-cancer was downloaded from [http://research-pub.gene.com/REdiscoverTEpaper/]. CRC Copy Number GISTIC2 level 4 data was downloaded from Broad GDAC Firehose [http://gdac.broadinstitute.org/runs/analyses_2016_01_28/data/COADREAD/20160128/]. The clinical data, gene program pathways were obtained from [https://xenabrowser.net/]. The TCR/BCR index scores and genetic changes were downloaded from the Supplementary table of the pan-cancer immune landscape paper (PMID: 29628290). Three cell line datasets were downloaded from [https://www.ncbi.nlm.nih.gov/geo/] under the accession number of (i) GSE5816, (ii) GSE80137, and (iii) GSE22250. 

## Author contributions

XZ and JWHW conceived this study. HF, KG, and JAB helped to collect the public data. XZ and JWHW conducted statistical analysis. HF, KG, and JAB helped to interpret the results. XZ and JWHW wrote the manuscript. Data used in this study is derived from TCGA database (https://cancergenome.nih.gov). Part of the data was analyzed on Cancer Genomic Cloud (CGC, http://www.cancergenomicscloud.org/). We thank the CGC team for helping to implement the TE analysis pipeline on CGC with pilot funds provided by Seven Bridges Genomics.

## Supplementary Materials

**Supplementary Methods**

**Supplementary Figures:**

**Supplementary Figure S1. Screening of candidate TEs. (A)** Schematic workflow of this study. DSS, disease free survival; OS, overall survival; DFI, disease free interval; PFI, progression free interval; ECM, extracellular matrix; VEGF, vascular endothelial growth factor; EMT, epithelial-mesenchymal transition; IPRES, innate anti-PD1 resistance signature; TMB, tumor mutation burden; CIMP, CpG island methylator phenotype; CNV, copy number variation; SNV, single-nucleotide variant; TLR, Toll-like receptors; ISGs, interferon-stimulated genes; DNMTi: DNA methyltransferase inhibitor. **(B)** Pie chart showing the fraction of 1,204 TEs at class level. **(C)** Venn diagram showing the overlaps of significant candidate TEs associated with four endpoints including OS, DFI, PFI, DSS. **(D-G)** Prognostic value of one representative TE (MSTA-int) with Kaplan-Meier survival analysis for OS **(D)**, DSS **(E)**, DFI **(F)** and PFI **(G)**. The hazard ratios (HR) and 95% CIs for pairwise comparisons in univariable analyses (log-rank test) are displayed in each Kaplan-Meier plot. **(H)** Density ridgeline plot showing FDR of univariable Cox regression analysis for each endpoint. Vertical line indicates the median value. **(I-J)** Forest plots showing univariable Cox regression analysis of TEs for four endpoints at family **(I)** and class **(J)** level, respectively. Solid dots represent the HR of death and open-ended horizontal lines represent the 95 % CIs. For TEs at family level, only those families significant with at least one endpoint were shown in **(I)**. Six main TEs at class level were shown. **(K-M)** Forest plots showing multivariable Cox regression analysis of six Retroposon at three endpoints including OS **(K)**, DSS **(L)** and PFI **(M)**. For each Retroposon at each endpoint, four clinical features were included for multivariable Cox regression analysis including age, gender, MSI status and AJCC stage. Solid dots represent the HR of death and open-ended horizontal lines represent the 95% CIs. **(N)** Bar plot showing the number of genes in 29 immune sets. **(O)** Density ridgeline plot showing FDR of Spearman’s correlation between TEs and immune sets. Vertical line indicates the median value. (P) histogram of nine TEs by number of TCGA cancer types where they are overexpressed.

**Supplementary Figure S2. Clinical and molecular comparison among TE clusters. (A)** Correlation matrix showing Spearman’s correlation coefficient among 9 TEs with each other. **(B)** Scatter plot of survival times (OS) against the prognostic factor (TE score). **(C-E)** Kaplan-Meier survival curves of the selected groups for K = 2 **(C)**, K = 3 **(D)** and K = 4 **(E)**. **(F)** Plot of the overall p-values against K with significance level α = 0.05. **(G)** Plot of the worst-pair p-values against K with significance level α = 0.05. **(H)** Heatmap showing the distribution of clinical and molecular features among four TE clusters. Each row represents one feature, column to each sample. P-value was calculated using chi-square test.

**Supplementary Figure S3. Comparison among TE cluster in terms of immune overdrive. (A)** Cell fraction of 10 cell types estimated using MCPCounter algorithm. P-value for each variable was calculated using Kruskal-Wallis test. For each variable, the median of normalized value in each cluster was shown. **(B)** Heatmap showing the expression profiles of Th-1 signatures compromised of 20 genes. Samples in each column was ordered by TE score with decreasing order from left to right. **(C)** Heatmap showing the expression profiles of MHC genes. Samples in each column was ordered by TE score with decreasing order from left to right. **(D)** Heatmap showing hotspot mutation profiles of 11 CRC drivers. Each row indicates one gene, each column indicates one sample. P-value was calculated using chi-square test by comparing between cluster 4 and three clusters combined. **(E)** CNV plot showing the GISTIC score among four TE clusters. **(F)** Heatmap showing the expression profiles of IPRES signatures compromised of 24 pathways. Each row indicates one pathway, each column indicates one sample. Sample in each column was ordered by TE score with decreasing order from left to right. **(G)** Gene set variation analysis of 39 gene program and canonical targetable pathways. 24 significant pathways were shown. P-value for each variable was calculated using Kruskal-Wallis test. For each variable, the median of normalized value in each cluster was shown. **(H)** Violin plot showing the comparison of CD8A expression amongst TE clusters. **(I)** Violin plot showing the comparison of CD274 expression amongst TE clusters. **(J)** Violin plot showing the difference of TE score among risk groups identified by Fakih et al (13) (****: p <= 0.0001). **(K)** t-SNE plot of TE expression profiles at subfamily level for 515 single cells. **(L)** Boxplot showing the proportion of reads mapping to TEs among cell types. P-value for each variable was calculated using Kruskal-Wallis test.

**Supplementary Figure S4. Construction of co-expression module using WGCNA. (A)** Clustering dendrogram of CRC samples. One sample (TCGA–AA–3947–01) was considered as outlier and was removed in downstream analysis. **(B-C)** Soft-thresholding power selection in WGCNA. Analysis of the scale-free fit index for individual soft-thresholding powers. Analysis of the mean connectivity for individual soft-thresholding powers. The power = 10 was chosen which is the lowest power for the curve that the scale-free topology fit index flat upon reaching a high value above 0.9 with a moderate mean connectivity. **(D)** Clustering dendrograms of genes included with dissimilarity based on topological overlap, together with assigned module colors. A total of 12 modules were identified and assigned into different colors. **(E-F)** Scatter plots of gene significance for TE score versus module membership in greenyellow **(E)** and brown **(F)** module, respectively. **(G)** Heatmap showing the topological overlap in WGCNA. Each row and column represents a gene, light color indicates low topological overlap, and progressively darker red indicates higher topological overlap. Darker squares along the diagonal represent modules. The gene dendrogram and module assignment are shown along the left and top. Heatmap on the right panel zooms into the brown and greenyellow modules.

**Supplementary Figure S5. Pan cancer analysis of TE score. (A)** Comparison of TE score across 24 cancer types. **(B)** Spearman’s correlation between TE score and GEP in pooled cancer samples (n = 6,554). **(C)** Heatmap showing the comparison of clinical and molecular features among four TE clusters in KIRC. Each row represents one feature, while each column represents one sample. P-value was calculated using the chi-square test. **(D-F)** Forest plots showing multivariable Cox regression analysis of TE cluster adjusted by clinical features for OS **(D)**, DSS **(E)** and PFI **(F)** in KIRC. All variable was set as categorial variable. Samples with age < 65 was set as age low group and ≥ 65 for high group. Solid dots represent the HR of death and open-ended horizontal lines represent the 95 % CIs. All P-values were calculated using Cox proportional hazards analysis (ns: p > 0.05, *: p <= 0.05, **: p <= 0.01, ***: p <= 0.001, ****: p <= 0.0001). **(G)** Gene set variation analysis showing fraction of 28 cell types in KIRC. **(H)** Gene set variation analysis showing immune infiltration signatures in KIRC. **(I)** TCR/BCR indexes comparison among TE clusters in KIRC. **(J)** Genetic changes comparison among TE clusters in KIRC. **(K)** Gene set variation analysis showing immune evasion signatures in KIRC. P-value for each variable was calculated using Kruskal-Wallis test. For each variable, the median of normalized value in each cluster was shown. **(L-R)** Representative expression of genes or signatures involved in immune response and RNA sensor signals in KIRC including RIG-I-like pathways **(L)**, APOBECs **(M)**, Oligoadenylate synthetases **(N)**, RNA sensors **(O)**, interferon-stimulated genes (ISGs) **(P)**, interferon secretion process **(Q)** and Toll-like receptors (TLRs) **(R)**. P-value for each variable was calculated using Kruskal-Wallis test. For each variable, the median of normalized value in each cluster was shown.

**Supplementary Figure S6. Correlation between TE score and global TE expression. (A)** Spearman’s correlation between TE score and the proportion of reads mapping to TEs in CRC. **(B)** Circle plot showing Spearman’s correlation between TE score and TE expression at five main class level (DNA, LINE, LTR, SINE and Retroposon) in CRC. **(C)** Spearman’s correlation between TE score and the proportion of reads mapping to TEs in scRNA-seq data of breast cancer. **(D)** Circle plot showing Spearman’s correlation between TE score and TE expression at the class level in KIRC.

**Supplementary tables:**

(The supplementary tables will be available upon publication of manuscript)

**Supplementary Table S1**. Results of screening TEs associated with survival.

**Supplementary Table S2**. Summary of clinical and molecular information from TCGA CRC samples analyzed

**Supplementary Table S3**. Gene signatures used in this study.

**Supplementary Table S4**. Overlapped significant pathways among DNMT inhibitor treated datasets and TE cluster.

**Supplementary Table S5**. Two module gene list derived from WGCNA.

**Supplementary Table S6**. Summary of clinical and molecular information from TCGA Pan-cancer samples analyzed

**Supplementary Table S7**. Univariable Cox regression analysis of TE score across 24 cancer types.

